# Prevalence of Cardiometabolic Disease Risk Factors in People living with HIV Initiating Anti-retroviral Therapy at a High-Volume HIV Clinic in Kampala, Uganda

**DOI:** 10.1101/2023.03.18.23287437

**Authors:** Willington Amutuhaire, Frank Mulindwa, Barbara Castelnuovo, Nele Brusselaers, Jean-Marc Schwarz, Mutebi Edrisa, Simon Dujanga, Robert A. Salata, George A. Yendewa

## Abstract

**Background:** Cardiometabolic diseases are a leading cause of HIV-related morbidity and mortality, yet routine screening is not undertaken in high burden countries. We aimed to assess the prevalence and risk factors of the metabolic syndrome (MetS) and its components in adult Ugandan people living with HIV (PLHIV) initiating dolutegravir-based antiretroviral therapy (ART).

**Methods:** We analyzed baseline data of PLHIV aged ≥ 18 years enrolled in the GLUMED (Glucose metabolism changes in Ugandan HIV patients on Dolutegravir) Study from January to October 2021. MetS was defined as having 3 or more of the following: abdominal obesity, hypertension (HTN), elevated fasting glucose, elevated triglycerides and low high-density lipoprotein cholesterol. Multiple logistic regression was used to assess associations between potential risk factors and MetS and its components.

**Results:** 309 PLHIV were analyzed (100% ART-naive, 59.2% female, median age 31 years and median CD4 count 318 cells/mm^3^). The prevalence of MetS was 13.9%. The most common cardiometabolic condition was dyslipidemia (93.6%), followed by abdominal obesity (34.0%), hyperglycemia (18.4%), and HTN (8.1%). In adjusted analysis, MetS was associated with age > 40 years (odds ratio 3.33, 95% confidence interval 1.45-7.67) and CD4 count > 200 cells/mm^3^ (3.79, 1.23-11.63). HTN was associated with age > 40 years (2.96, 1.32-6.64), and dyslipidemia was associated with urban residence (4.99, 1.35-18.53).

**Conclusion:** Cardiometabolic risk factors were common in this young Ugandan cohort of PLHIV initiating dolutegravir-based ART, underscoring the need for programmatic implementation of surveillance and management of comorbidities in Uganda and similar settings.

**Key points:** In Ugandan HIV patients initiating antiretroviral treatment, the prevalence of cardiometabolic risk factors was substantial: metabolic syndrome 13.9%, dyslipidemia 93.6%, abdominal obesity 34.0%, hyperglycemia 18.4%, and hypertension 8.1%. This highlights the need for early screening and management of comorbidities.

## BACKGROUND

The HIV epidemic remains a major public health problem, with 38 million people infected globally in 2021[1]. Despite this, increased access to potent combination anti-retroviral therapy (ART) has substantially lowered HIV-related morbidity and mortality and prolonged the life expectancy of people living with HIV (PLHIV). This is of utmost relevance in sub-Saharan Africa (SSA) where the global burden of HIV is highest[2], [3]. Paradoxically, this has resulted in an increase in the burden of non-communicable diseases (NCDs) among PLHIV[3]–[6]. Globally, NCDs cause 41 million deaths annually, with cardiovascular diseases (CVDs) accounting for nearly half (44%) of such deaths[7]. PLHIV are at a two-fold higher risk of experiencing myocardial infarction, strokes and other CVDs due to multiple factors, including having a higher prevalence of traditional CVD risk factors at baseline, ART-associated toxicities, HIV-induced immune activation and chronic systemic inflammation[8], [9]. With recent modelling studies projecting an excess increase in NCDs among PLHIV[10], [11], the Joint United Nations Programme on HIV/AIDS (UNAIDS) is advocating for increased surveillance and integration of NCD care in HIV programs[12]–[14].

Since 2016, the World Health Organization (WHO) has recommended the use of integrase strand transfer inhibitors (INSTI) as the preferred first- and second-line ART[15], [16]. This followed multiple reports of increasing primary HIV drug resistance (HIVDR) to non-nucleoside reverse transcriptase inhibitors (NNRTIs), with many high burden countries reporting NNRTI HIVDR prevalence rates exceeding the 10% recommended threshold[17], [18]. Subsequently, most countries in SSA have moved to adopt dolutegravir (DTG)-based regimens as first-line ART, due to its high barrier to resistance, tolerability, and less drug-drug interactions [19]–[21]. Despite their tolerability, however, INSTIs have consistently been linked to excessive weight gain, particularly among women, older patients, and people of Black or African ancestry[22]–[24]. Additionally, analyses for insulin resistance trajectories from multiple clinical trial data, e.g., SPRING-2 [23], SAILING [24], and VIKING-3 described worsening insulin resistance in patients post switch to INSTI-based AR[25]. Similarly, multiple case reports describing real world experiences with INSTI use have documented accelerated hyperglycemia in patients post switch to INSTI-based ART [26]–[28]. In one such a report from Uganda, there was an observed incidence rate of 4.7 cases of hyperglycemia per 1000 patients in a median duration of 4 months after initiating DGT-based ART, with the majority (15 of 16 cases) presenting with diabetic ketoacidosis[26]. Consequently, the Ugandan Ministry of Health HIV treatment guidelines halted the use of DTG in patients known to have diabetes or at high risk of cardiometabolic diseases[29], [30].

Given the well described metabolic complications associated with INTSIs, their widespread adoption as first- and second-line ART in SSA is likely to significantly exacerbate the growing problem of NCDs among PLHIV in this region. It is therefore essential that HIV treatment programs in SSA implement effective NCD control measures focused on early screening and prevention of comorbidities. However, there are limited studies to inform evidence-based public health policy and clinical practice. In this study, we aimed to assess the prevalence and risk factors associated with the metabolic syndrome (MetS) and its components in a cohort of treatment naïve PLHIV starting ART at a high-volume HIV clinic in Kampala, Uganda.

## METHODS

### Study design and setting

The GLUMED (Glucose metabolism changes in Ugandan HIV patients on Dolutegravir) Study was a prospective cohort study that enrolled newly diagnosed PLHIV prior to ART initiation between January and September 2021 at Kisenyi Health Center HIV Clinic in Kampala, the capital and largest city in Uganda. The primary objective of the study was to describe changes in glucose metabolism in Ugandan PLHIV at study entry and 48 weeks after initiating DTG-based ART. The Kisenyi Health Center HIV Clinic is one of the busiest in the country, with about 12,000 active PLHIV enrolled in routine care. We analyzed baseline sociodemographic and clinical data on patients who were sequentially enrolled in the study.

### Study participants

Between January and October 2021, ART naïve PLHIV aged 18 years or older starting DTG-based ART were screened for study inclusion. HIV status was determined using the rapid test by SD Bioline HIV-1/2 3.0 (Standard Diagnostics Inc, Suwon, Korea). Pregnant women and very sick patients deemed unable to undergo a 2 hour -75g oral glucose tolerance test (OGTT) were excluded. Criteria for exclusion during follow up included: pregnancy, development of T2DM, non-adherence to study protocols and poor adherence to ART.

Of the 435 PWH screened, 310 patients (71%) consented for oral glucose tolerance test evaluation for study enrollment. One patient was lost to follow up, leaving 309 patients that provided data for analysis. (Figure 1)

**Figure 1.**
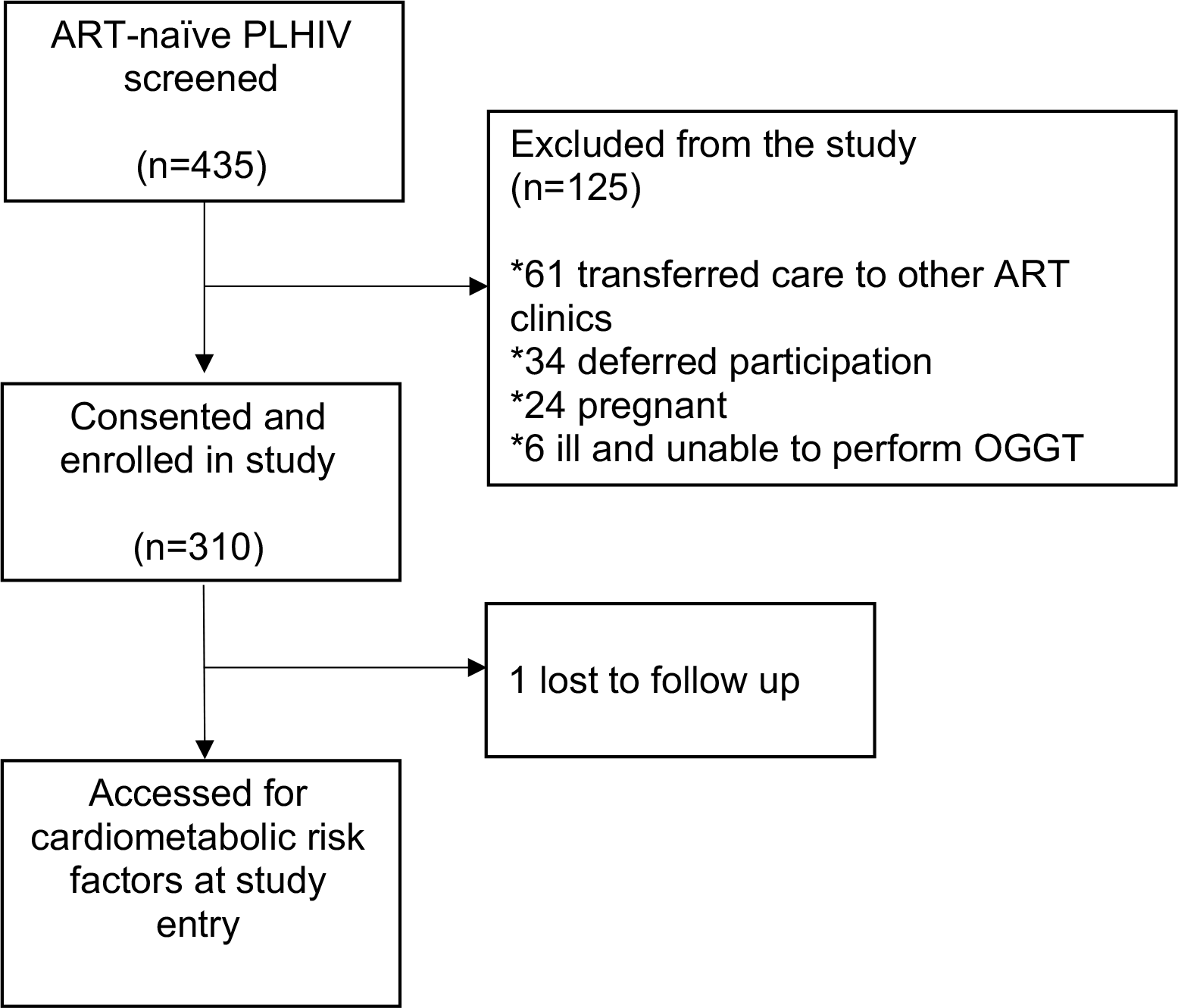
Enrollment schema for the GLUMED study.

### Study processes

After consenting, patients were scheduled for review in 24 to 48 hours after an overnight fast of 8-12 hours. Peripheral venous cannulae were placed and 5 ml of fasting venous blood was collected in plain 5ml BD Vacutainers®. Patients were then given 300 ml of 75 gram-glucose solution to be taken within 5 minutes. Repeat 5 ml blood samples were collected at: 30, 60, 90 and 120 minutes from the time of ingestion of the glucose solution. Blood glucose was measured at all time points immediately after sample collection (before transfer to vacutainers) using ACCU-CHECK™ glucometers (Roche Diagnostics, Manheim, Germany) [31]. After centrifugation of the whole venous blood samples, serum was collected and stored at -80^0^C for batched serum lipids, creatinine, insulin and C-peptide analysis. Patients found to have a normal 2-hour 75g OGTT (fasting blood glucose <126mg/dl and 2-hour glucose <200mg/dl) were enrolled for 48-week follow up on tenofovir/ lamivudine/ dolutegravir as recommended by the Uganda National HIV treatment guidelines. [32]

Baseline demographic, clinical and social data were collected which included: age, sex, CD4 count (determined by Alere Pima Analyzer (Abbott Diagnostics, Jena, Germany), body mass index (BMI), level of education, area of residence, blood pressure, waist circumference, tuberculosis (TB) status, smoking status, physical activity measured by the Global Physical Activity Questionnaire (GPAQ), alcohol consumption measured by the Alcohol Use Disorders Identification Test (AUDIT), serum creatine and serum lipid profiles (Cobas 6000^®^ from Roche Diagnostics).

### Assessments and variable definitions

Risk of harmful alcohol use was assessed using the standardized AUDIT questionnaire[33]. Level of physical activity was reported as meeting or not meeting the WHO Global physical activity questionnaire (GPAQ) thresholds [30].

Waist circumference (WC) measurements were used to assess abdominal obesity and risk of cardiometabolic disease was stratified according to the WHO criteria[34], as follows: (1) not at risk: < 95 cm (males) and < 81 cm (females); (2) at increased risk: males ≥ 95 to ≤102 cm and females ≥ 81 to ≤ 88 cm; and (3) at substantially increased risk: ≥ 103 cm (males) and ≥ 89 cm (females). Body mass index (BMI) was classified based on the CDC[35] as follows (1) underweight: < 18.5 kg/m^2^, (2) normal: 18.5 -24.9 kg/m^2^, (3) overweight: 25-29.9 kg/m^2^, and obese: ≥30 kg/m^2^ for obese[35]. Hypertension was assessed as the average of two readings or being on antihypertensive treatment and was categorized in accordance with the Joint National Committee 8 (JNC-8) guidelines[36], as follows: (1) normal: systolic blood pressure (SBP) < 120 mmHg and diastolic blood pressure (DBP) < 80 mmHg; (2) pre-hypertension: SBP 120-139 mmHg or DBP 80-89 mmHg; and (3) hypertension (HTN): SBP ≥ 140 mmHg or DBP ≥ 90 mmHg.

Blood glucose level was categorized in accordance with American Diabetes Association criteria[37], as follows: (1) normal: fasting glucose < 100 mg/dL or 2-hour OGGT < 140 mg/dL; (2) prediabetes: fasting glucose 100-125 mg/dL or 2-hour glucose 140-199 mg/dL; and (3) diabetes: fasting glucose ≥ 126 mg/dL or 2-hour or glucose ≥ 200 mg/dL. Dyslipidemia was defined as having any one of the following[38]: total cholesterol (TC) > 200 mg/dL, high-density lipoprotein cholesterol (HDL-C) < 40 mg/dL (males) or < 50 mg/dL (females), low-density lipoprotein cholesterol (LDL-C) > 100 mg/dL and triglycerides (TG) > 150 mg/dL. Glomerular filtration rate (eGFR) was estimated using the Modification of Diet in Renal Disease (MDRD) Study equation[39], with renal impairment defined as eGFR < 60 mL/min/1.73 m^2^. Laboratory-based 10-year CVD (Framingham) risk scores were calculated for participants aged ≥ 30 years (170/309 (55%)) and classified as follows: low risk: < 10%, intermediate risk: 10-20% and high risk; >20%[40]. Lastly, we used a harmonized definition of the MetS based on diagnostic criteria proposed by the International Diabetes Federation (IDF) Task Force on Epidemiology and Prevention, the American Heart Association/National Heart, Lung, and Blood Institute (AHA/NHLBI) and the World Health Organization (WHO)[41], which requires having at least 3 of the following 5 criteria: (1) WC > 94 cm (men) or >81 cm women); (2) TG ≥ 150 mg/dL; (3) HDL-C < 40 mg/dL (males) or < 50 mg/dL(females); (4) fasting glucose ≥ 100 mg//dL; and (5) SBP ≥ 130 mm Hg or DBP ≥ 85 mm Hg.

### Statistical analysis

Statistical analyses were performed using SPSS Version 29.0 (Armonk, NY; IBM Corp). Categorical variables were reported as frequencies (percentages) and associations assessed using Pearson’s chi-square or Fisher’s exact tests. Continuous variables were presented as medians (interquartile ranges, IQR) and associations assessed using the non-parametric independent samples Mann–Whitney U-test. A logistic regression model was used to identify associations between MetS and each of its components (i.e., HTN, dyslipidemia, and hyperglycemia) and known or potential associated risk factors. Only variables that attained a p-value of < 0.2 in the univariate analysis were included in the multivariate regression model. Associations were reported as crude (OR) and adjusted odds ratios (AOR) with 95% confidence intervals (CI), p < 0.05 considered statistically significant.

### Patient Consent Statement

Ethical approval to perform the study was obtained from The AIDS Support Organization (TASO) Institutional Review Board (registration number: TASOREC/051/2020-UG-REC-009) and the Uganda National Council for Science and Technology (Registration number: HS1032ES). All participants signed informed consent prior to enrolment in the study.

## RESULTS

### Baseline characteristics of study participants

A total of 309 PWH were enrolled in the study and the majority (59.2%, 183/309) were female. The median age was 31 years (IQR 27-39), with males being older than females (median ages 34 vs 30 years, p < 0.001). Most were single (57.6%, 178/309), had attained primary education or higher (98.7%, 305/309), were Christian (79.3%, 245/309), resided in urban areas (92.2%, 285/309) and were employed (81.9%, 253/309). Of the 259 PWH with available data, the median CD4 count was 318 cells/mm^3^ (Interquartile range (IQR): 163-550) at enrollment. (Table 1)

**Table 1.** Baseline characteristics of PWH by sex before ART initiation (N=309)

| Characteristics | All PWH | Male | Female | p-Value |
| --- | --- | --- | --- | --- |
| **Total** | 309 | 126 | 183 |  |
| **Age, years** |  |  |  |  |
| Median (IQR) | 31 (27-39) | 34 (28-40) | 30 (26-37) | **<0.001** |
| < 30 | 122 (39.5) | 35 (27.8) | 87 (47.5) | **0.002** |
| 30-39 | 121 (39.2) | 57 (45.2) | 64 (35.0) |  |
| 40-49 | 57 (18.4) | 31 (24.6) | 26 (14.2) |  |
| ≥50 | 9 (2.9) | 3 (2.4) | 6 (3.3) |  |
| **Relationship status** |  |  |  |  |
| Single | 178 (57.6) | 62 (49.2) | 116 (63.4) | **0.013** |
| Married | 131 (42.4) | 64 (50.8) | 67 (36.6) |  |
| **Education** |  |  |  |  |
| None | 4 (1.3) | 0 | 4 (2.2) | **0.015** |
| Primary | 166 (53.7) | 74 (58.7) | 92 (50.3) |  |
| Secondary | 126 (40.8) | 43 (34.2) | 83 (45.4) |  |
| Tertiary | 13 (4.2) | 9 (7.1) | 4 (2.1) |  |
| **Religion** |  |  |  |  |
| Christian | 245 (79.3) | 102 (81.0) | 143 (78.1) | 0.549 |
| Muslim | 64 (20.7) | 24 (19.0) | 40 (21.9) |  |
| **Residence** |  |  |  |  |
| Urban | 285 (92.2) | 120 (95.2) | 165 (90.2) | 0.101 |
| Rural | 24 (7.8) | 6 (4.8) | 18 (9.8) |  |
| **Employment** |  |  |  |  |
| Yes | 253 (81.9) | 120 (95.2) | 133 (72.7) | **<0.001** |
| No | 56 (18.1) | 6 (4.8) | 50 (27.3) |  |
| **Baseline CD4 count, cells/mm<sup>3</sup>**<br>(n=259) |  |  |  |  |
| Median (IQR) | 318 (163-550) | 251 (123-382) | 399 (238-651) | **<0.001** |
| ≤99 | 36 (13.9) | 21 (20.4) | 15 (9.6) | **<0.001** |
| 100-199 | 37 (14.3) | 21 (20.4) | 16 (10.3) |  |
| 200-349 | 66 (25.5) | 31 (30.1) | 35 (22.4) |  |
| ≥350 | 120 (46.3) | 30 (29.1) | 90 (57.7) |  |

### Prevalence of cardiometabolic risk factors

The overall proportion of patients with a history of smoking and harmful alcohol use (AUDIT ≥ 16) was low at 4.9% (15/309) and 6.5% (20/309), respectively. The majority (78.0%, 241/309) met the WHO daily goals for physical activity.

The median BMI was 22.2 kg/m^2^, with women more likely to have higher median BMIs compared with their male counterparts (p < 0.001). Overall, 21.7% (67/30) were classified as overweight (pre-obesity, BMI 25-29.9 kg/m^2^), while 8.1% (25/309) were classified as obese (BMI ≥ 30 kg/m^2^). Based on the WC measurements, 18.4% (57/309) of participants were at increased risk cardiometabolic disease, while 15.5% (48/309) were at a substantially higher risk. There were sex differences observed, with women more likely to have higher WC measurements (p < 0.001).

A substantial proportion of study participants had prehypertension (16.8%, 52/309), while 8.1% (25/309) had HTN. The prevalence of overt diabetes was low at 0.3% (1/309); however, 18.1% (56/309) had prediabetes. Of those with available testing for kidney function, the median serum creatinine was 0.80 mg/dL, with a small proportion of study participants (1.4%, 4/282) having impaired renal function (eGFR < 60 ml/min/1.73m^2^).

About 93.6% (264/282) of study participants had dyslipidemia. The prevalence of hypercholesterolemia (TC > 200 mg/dL) was low at 5.3% (15/282). Of the individual components of the lipid panel, 88.3% (249/282) had low HDL-c (HDL-c < 40 mg/dL), 20.9% (59/282) had elevated LDL-c (LDL-c > 100 mg/dL), while 13.1% (37/282) had elevated triglycerides (TG > 150 mg/dL). More women had derangements in the individual factions of the lipid panel compared with their male counterparts (all p < 0.001).

Overall, 13.9% (43/309) of study participants met the criteria for the metabolic syndrome, without sex differences (p=0.129). Despite this, only a small proportion (1.8%, 3/170) had a 10-year CVD risk > 10%. (Table 2)

**Table 2.** Cardiometabolic risk factors by sex in PWH prior to ART initiation

| Risk factors | All PWH | Male | Female | p-Value |
| --- | --- | --- | --- | --- |
| **Total** | 309 | 126 | 183 |  |
| **Active or past smoking** |  |  |  |  |
| Yes | 15 (4.9) | 9 (7.1) | 6 (3.3) | 0.120 |
| No | 294 (95.1) | 117 (92.9) | 177 (96.7) |  |
| **Alcohol use (AUDIT scores)** |  |  |  |  |
| 0-7 (low-risk) | 263 (85.1) | 97 (77.0) | 166 (90.7) | **0.011** |
| 8-15 (at risk) | 26 (8.4) | 16 (12.7) | 10 (5.5) |  |
| 16-19 (high risk) | 12 (3.9) | 8 (6.3) | 4 (2.2) |  |
| ≥20 (addiction) | 8 (2.6) | 5 (4.0) | 3 (1.6) |  |
| **Level of physical activity** |  |  |  |  |
| <600 MET (does not meet WHO goals) | 68 (22.0) | 22 (17.5) | 46 (25.1) | 0.109 |
| ≥600 MET (meets WHO goals) | 241 (78.0) | 104 (82.5) | 137 (74.9) |  |
| **Body mass index, kg/m<sup>2</sup>** |  |  |  |  |
| Median (IQR) | 22.2 (19.7-25.6) | 21.1 (19.2-22.9) | 23.4 (20.7-27.6) | **<0.001** |
| <18.5 (underweight) | 32 (10.4) | 16 (12.7) | 16 (8.78) | **<0.001** |
| 18.5-24.9 (normal) | 185 (59.9) | 96 (76.2) | 89 (48.6) |  |
| 25.0-29.9 (pre-obesity) | 67(21.7) | 10 (7.9) | 57 (31.1) |  |
| ≥30 (obese) | 25 (8.1) | 4 (3.2) | 21 (11.5) |  |
| **Waist circumference, cm** |  |  |  |  |
| Median (IQR) | 79.0 (75.0-86.0) | 79.0 (75.0-83.0) | 81.0 (74.0-88.0) | 0.153 |
| Not at risk (males: <95 cm and females: <81 cm) | 204 (66.0) | 115 (91.3) | 89 (48.6) | **<0.001** |
| Increased risk (males: ≥95 to ≤102 cm and females: ≥81 to ≤88 cm) | 57 (18.4) | 8 (6.3) | 49 (26.8) |  |
| Substantially increased risk (males: ≥ 103 cm and females: ≥89 cm) | 48 (15.5) | 3 (2.4) | 45 (24.6) |  |
| **Blood pressure, mmHG** |  |  |  |  |
| Normal (<120/80 mmHg) | 232 (75.1) | 92 (73.0) | 140 (76.5) | 0.115 |
| Pre-hypertension (120-139/80-89 mmHg) | 52 (16.8) | 19 (15.1) | 33 (18.0) |  |
| Hypertension (≥ 140/90 mmHg) | 25 (8.1) | 15 (11.9) | 10 (5.5) |  |
| **Laboratory-based CVD risk score \***<br>(n=170) |  |  |  |  |
| <5% | 149 (87.6) | 67 (79.8) | 82 (95.3) | **0.007** |
| 5-10% | 18 (10.6) | 15 (17.9) | 3 (3.5) |  |
| >10% | 3 (1.8) | 2 (2.4) | 1 (1.2) |  |
| **Blood glucose, mg/dL** |  |  |  |  |
| Normal (fasting < 100 mg/dL or 2-hour OGGT <140 mg/dL) | 252 (81.6) | 98 (77.8) | 154 (84.2) | 0.150 |
| Prediabetes (fasting 100-125 mg/dL or 2-hour glucose 140-199 mg/dL) | 56 (18.1) | 27 (21.4) | 29 (15.8) |  |
| Diabetes (fasting ≥ 126 mg/dL or 2-hour or glucose ≥ 200 mg/dL) | 1 (0.3) | 1 (0.8) | 0 (0) |  |
| **Serum creatinine, mg/dL (n=281)** |  |  |  |  |
| Median (IQR) | 0.80 (0.68-0.93) | 0.89 (0.77-1.01) | 0.76 (0.64-0.84) | **<0.001** |
| **eGFR, ml/min/1.73m<sup>2</sup> (n=281)** |  |  |  |  |
| Median, IQR | 111 (96-129) | 118 (99-135) | 106 (94-122) | 0.231 |
| ≥90 (Stage 1) | 230 (81.9) | 101 (86.3) | 129 (78.7) |  |
| 60-89 (Stage 2) | 47 (16.7) | 14 (12.0) | 33 (20.1) |  |
| 45-59 (Stage 3a) | 2 (0.7) | 1 (0.9) | 1 (0.6) |  |
| 30-44 (Stage 3b) | 2 (0.7) | 1 (0.9) | 1 (0.6) |  |
| 29-15 (Stage 4) | 0 (0) | 0 (0) | 0 (0) |  |
| <15 (Stage 5) | 0 (0) | 0 (0) | 0 (0) |  |
| **Total cholesterol (TC), mg/dL (n=282)** |  |  |  |  |
| Median, IQR | 132 (112-157) | 125 (106-147) | 138 (115-163) | **0.002** |
| TC > 200 mg/dL | 15 (5.3) | 4 (3.4) | 11 (6.7) | 0.288 |
| **HDL-C, mg/dL (n=282)** |  |  |  |  |
| Median, IQR | 31 (24-38) | 27 (22-36) | 34 (26-41) | **<0.001** |
| HDL-C < 40 mg/dL | 249 (88.3) | 101 (86.3) | 148 (89.7) |  |
| **LDL-C, mg/dL (n=282)** |  |  |  |  |
| Median, IQR | 73 (53-93) | 66 (50-88) | 78 (56-100) | **0.007** |
| LDL-C > 100 mg/dL | 59 (20.9) | 14 (12.0) | 45 (27.3) | **0.002** |
| **Triglycerides (TG) mg/dL (n=282)** |  |  |  |  |
| Median, IQR | 89 (62-118) | 93 (70-124) | 86 (59-116) | 0.058 |
| TG > 150 mg/dL | 37 (13.1) | 19 (16.2) | 18 (10.9) | 0.191 |
| **Dyslipidemia<sup>a</sup> (any 1 of the following: elevated TC<sup>b</sup>, low HDL-C<sup>c</sup>, elevated LDL-C<sup>d</sup> or elevated TG<sup>e</sup>) (n=282)** |  |  |  |  |
| Yes | 264 (93.6) | 106 (90.6) | 158 (95.8) | 0.081 |
| No | 18 (6.4) | 11 (9.4) | 7 (4.2) |  |
| **Metabolic syndrome (any 3 of the following: central obesity<sup>f</sup>, elevated fasting blood glucose<sup>g</sup>, HTN<sup>g</sup>, low HDL-C<sup>c</sup> or elevated TG<sup>e</sup>)** |  |  |  |  |
| Yes | 43 (13.9) | 13 (10.3) | 30 (16.4) | 0.129 |
| No | 266 (86.1) | 113 (89.7) | 153 (3.6) |  |

\*- calculated by the laboratory-based Framingham risk score, b- Total cholesterol, c- High density lipoproteins, d- Low density lipoproteins, e- Triglycerides, f- , g-, h

### Associated Risk Factors of MetS and its components

In adjusted multivariate logistic regression analysis, MetS was significantly associated with age > 40 years (Adjusted Odds Ratio (AOR) 3.33, 95% confidence interval (CI) 1.45-7.67) and CD4 count > 200 cells/mm^3^ (AOR 3.79, 95% CI 1.23-11.63). Similarly, hypertension was significantly associated with age > 40 years (AOR 2.96, 95% CI 1.32-6.64). Dyslipidemia was predicted by residence in an urban area (AOR 4.99, 95% CI 1.35-18.53); however, there were no independent predictors for hyperglycemia (Table 3).

**Table 3.** Predictors of the MetS and its components in PWH prior to ART initiation

| Risk factors | Metabolic syndrome |  | Hypertension |  | Dyslipidemia |  | Hyperglycemia |  |
| --- | --- | --- | --- | --- | --- | --- | --- | --- |
|  | OR<br>(95%CI) | AOR<br>(95% CI) | OR<br>(95%CI) | AOR<br>(95% CI) | OR<br>(95%CI) | AOR<br>(95% CI) | OR<br>(95%CI) | AOR<br>(95% CI) |
| **Gender** |  |  |  |  |  |  |  |  |
| Male | Ref | Ref | Ref | Ref | Ref | Ref | Ref |  |
| Female | **1.70 (0.85-3.41)** | 1.44 (0.64-3.25) | **0.61 (0.33-1.11)** | 0.58 (0.24-1.41) | **2.34 (0.88-6.25)** | 1.56 (0.51-4.77) | 0.76 (0.36-1.58) |  |
| **Age, years** |  |  |  |  |  |  |  |  |
| < 40 | Ref | Ref | Ref | Ref | Ref |  | Ref | Ref |
| ≥ 40 | **3.27 (1.65-6.47)** | **3.33 (1.45-7.67)\*\*** | **2.67 (1.39-5.10)** | **2.96 (1.32-6.64)\*\*\*** | 0.96 (0.31-3.04) |  | **1.79 (0.80-4.00)** | 1.59 (0.70-3.59) |
| **Relationship status** |  |  |  |  |  |  |  |  |
| Single | Ref |  | Ref |  | Ref |  | Ref |  |
| Married | 1.35 (0.71-2.58) |  | 1.25 (0.69-2.29) |  | 0.57 (0.22-1.49) |  | 0.92 (0.44-1.94) |  |
| **Education** |  |  |  |  |  |  |  |  |
| Primary or less | Ref |  | Ref | Ref | Ref |  | Ref |  |
| Secondary/tertiary | 1.07 (0.56-2.05) |  | **1.62 (0.87-3.02)** | 1.61 (0.77-3.33) | 1.01 (0.39-2.64) |  | 1.41 (0.66-2.99) |  |
| **Religion** |  |  |  |  |  |  |  |  |
| Christian | 1.40 (0.59-3.32) |  | 1.49 (0.66-3.35) |  | 0.46 (0.10-2.08) |  | 0.76 (0.32-1.78) |  |
| Muslim | Ref |  | Ref |  | Ref |  | Ref |  |
| **Residence** |  |  |  |  |  |  |  |  |
| Urban | 1.14 (0.33-4.01) |  | 0.73 (0.26-2.06) |  | **4.43 (1.31-15.01)** | **4.99 (1.35-18.53)\*\*\*** | 0.79 (0.22-2.82) |  |
| Rural | Ref |  | Ref |  | Ref | Ref | Ref |  |
| **Employment** |  |  |  |  |  |  |  |  |
| Yes | 1.43 (0.57-3.57) |  | **2.27 (0.86-5.99)** | 1.41 (0.49-4.08) | **0.25 (0.03-1.94)** | 0.31 (0.04-2.62) | **3.63 (0.84-15.67)** | 3.55 (0.81-15.51) |
| No | Ref |  | Ref | Ref | Ref | Ref | Ref | Ref |
| **Smoking** |  |  |  |  |  |  |  |  |
| Yes | 1.59 (0.43-5.87) |  | 1.28 (0.35-4.71) |  | 0.74 (0.09-6.07) |  | - |  |
| No | Ref |  | Ref |  | Ref |  |  |  |
| **Alcohol use (AUDIT scores)** |  |  |  |  |  |  |  |  |
| Low-moderate | Ref |  | Ref |  | Ref |  | Ref |  |
| High risk | 0.67 (0.15-3.01) |  | 0.54 (0.12-2.42) |  | 0.59 (0.13-2.75) |  | 0.96 (0.21-4.34) |  |
| **Physical activity** |  |  |  |  |  |  |  |  |
| High | Ref |  | Ref |  | Ref |  | Ref |  |
| Low | 0.92 (0.43-1.98) |  | 0.90 (0.44-1.83) |  | 1.44 (0.40-5.14) |  | 1.45 (0.64-3.29) |  |
| **Body mass index, kg/m<sup>2</sup>** |  |  |  |  |  |  |  |  |
| <25 | - | - | Ref | Ref | Ref | Ref | Ref | Ref |
| ≥25 | - | - | **2.25 (1.21-4.17)** | 2.94 (0.83-10.43) | **7.93 (1.04-60.61)** | 6.29 (0.48-81.57) | **1.72 (0.81-3.68)** | 1.78 (0.83-3.81) |
| **Waist circumference** |  |  |  |  |  |  |  |  |
| Normal | - | - | Ref | Ref | Ref | Ref | Ref |  |
| Increased | - | - | **1.57 (0.85-2.90)** | 1.16 (0.32-4.35) | **4.63 (1.05-20.64)** | 1.03 (0.14-7.45) | 1.00 (0.46-2.17) |  |
| **CD count cells/mm<sup>3</sup>** |  |  |  |  |  |  |  |  |
| <200 | Ref | Ref | Ref | Ref | Ref |  | Ref |  |
| ≥200 | **3.32 (1.13-9.78)** | **3.79 (1.23-11.63)\*\*\*** | **2.50 (1.00-6.23)** | **2.90 (1.09-7.73)\*\*** | 0.62(0.17-2.26) |  | 1.01 (0.40-2.53) |  |

Variables that were found to be significant, <.20 in univariable and <.05 in multivariable, are marked in bold.
Significance p-levels:
\*\*p <0.05.
\*\*\*p <0.01.

## DISCUSSION

In this study of treatment-naïve adult PLHIV initiating ART in Uganda, we estimated the baseline prevalence of MetS and key CVD risk factors with the aim of enhancing early screening and timely prevention of NCDs among Ugandan PLHIV. In mixed population studies, HIV-positive status has consistently conferred a higher risk of MetS and its components[42]. This underscores the necessity for pre-emptive screening for MetS both at baseline and prospectively while on ART.

Despite being a relatively young cohort (median age 31 years), the prevalence of MetS was 13.9% (i.e., about 1 in 7 patients) in our study. While several studies have assessed the prevalence of MetS among PLHIV in SSA, few have incorporated metabolic profiling at ART initiation. Estimates of the prevalence of MetS among PLHIV in SSA have generally been reported to be higher than our estimate, ranging from 20% to 58% [42]– [46]. This is likely a reflection of the fact that in most of these studies, participants were older and had a history of exposure to early ART-era protease inhibitors (PIs) and NRTIs, which were associated with higher rates of lipid and glucose dysmetabolism [47]–[50].

Dyslipidemia was the most common CVD risk factor in our cohort, affecting about 93.6% of patients. Of these, low HDL-C was by far the most important risk factor and was present in 88.6% (or nearly 9 in 10 patients). Few studies from SSA have documented the lipid profile of PLHIV pre-ART; most reports have focused largely on ART-exposed individuals. In five cross-sectional studies of ART-exposed PLHIV from Malawi, Nigeria, Ethiopia and Cameroon, the prevalence of low HDL-C, elevated LDL-C, TG and TC were 34-63%, 17-21%, 9% and 11-24%, respectively, which were lower than our findings [51]– [54]. In these and other studies from SSA, advanced age, duration on ART, increased waist-to-hip ratio, high BMI and a sedentary lifestyle have been identified as risk factors of dyslipidemia [51]–[59]. We did not detect any statistically significant associations between these traditional risk factors and dyslipidemia in our cohort; however urban residence conferred a 5-fold higher risk of dyslipidemia. Plausible explanations for this finding include a higher prevalence of traditional CVD risk factors among urban dweller compared to their rural counterparts such as such as unhealthy dietary habits, a more sedentary lifestyle and greater levels of exposure to environmental pollutants among urban dwellers (e.g., emissions from fossil fuel combustion and industrial processes). Careful studies are needed to confirm these hypotheses.

Hypertension is a common comorbidity among PLHIV and was estimated to affect 24% of PLHIV globally in recent meta-analysis by Bigna *et al* [60]. In this and other studies, geographic variations in the prevalence of hypertension among PLHIV have been reported, with countries in SSA being disproportionately affected [61]-[67]. Despite the prevalence of hypertension being lower in our study (8.1%), a substantial proportion (17%) were pre-hypertensive and at risk of progressing to hypertension. Similarly, the prevalence of diabetes mellitus in our study was very low (0.3%), while pre-diabetes was higher (18.1%). The link between diabetes and HIV is controversial, with some studies suggesting that HIV is an independent risk factor of diabetes [68], [69], while others have failed to establish a clear link [70], [71]. However, as previously discussed, PLHIV exposed to ART (especially INTSI-based regimens) are at risk of metabolic complications including weight gain, increased insulin resistance and the metabolic syndrome [22]-[30]. Thus, the identification of pre-hypertension and prediabetes prior to ART exposure offers opportunities to intervene early with non-pharmacological risk-reduction strategies (e.g., smoking cessation, increased physical exercise, weight loss and reduced salt-intake) to lower overall CVD risk.

Of the traditional CVD risk factors that were assessed in our study, increasing age (≥40 years) and CD4 count(≥ 200 cells/mm^3^) were associated with a higher risk of both MetS and hypertension, respectively. The latter finding is consistent with previous studies which have shown that higher CD4 cell count is an independent predictor of the development of MetS, attributable largely to the expected improvement in nutritional intake, weight gain and energy conservation associated with initiation ART [72]-[74]. However, our cohort was entirely ART-naïve. It remains unclear what mechanisms are at play linking degree of immunosuppression and CVD risk stratification in ART-naïve patients. Further studies are needed to explore the potential underlying causes of this finding.

We used laboratory-based Framingham scores to estimate 10-year CVD risk for study participants above 30 years (50% of enrolled participants) and determined that the majority had low 10-year CVD risk. In three studies from Cameroon and South Africa, 50 to 93% of PLHIV had a 10-year CVD risk of < 5% [75]-[77], compared with 87% of patients in our study. Most CVD risk calculators, including the Framingham score that we used in our study tend to underestimate the 10-year CVD risk, as they do not take into account other patient characteristics such as the inherent effect of HIV itself, systemic inflammation, as well as infectious and non-communicable comorbidities [78]. This may explain the possible underestimation of risk in the cited studies above, involving much older patients on ART, a known risk factor to both micro- and macro-vascular disease[47]. Despite the reported low 10-year risk of CVD in our study population, we demonstrated that this is a population with high frequencies of individual CVD risk factors, and therefore hypothesize that their actual CVD risk may be much higher than the calculated risk.

Another noteworthy finding of our study was that although women had a higher prevalence of individual CVD risk factors (e.g., higher BMI, waist circumference, higher LDL and TG) compared with their male counterparts, there was no sex difference in the overall prevalence of MetS. Sex differences are known to influence the occurrence of MetS across different age groups in the general population [79]. A large part of this difference is attributable to the cardioprotective effects of estrogen and other hormones on glucose and lipid metabolism in women [79] [75]; the reduction in hormonal influence in post-menopausal women is implicated in the observed increase in seen the prevalence MetS and CVD risk in age-matched controls [79] [75]. Similar to our study, most studies which have reported on the prevalence of MetS in SSA have been in ART-exposed PLHIV and have not shown a sex differential effect [44], [80].

Our study had several limitations worthy of discussion. Firstly, the definition of MetS and its key components are based on models developed in high income countries and may not be applicable to populations in SSA, which may exhibit significant phenotypic and genotypic differences. Secondly, this was a single center study in an urban setting, hence the findings may not be generalizable to the wider Ugandan PLHIV population. Thirdly, based on study inclusion criteria, sicker patients were excluded due to their inability to undergo a 2-hour OGTT, which would have resulted in under-estimation of the prevalence of MetS and its components. Despite these limitations, while most studies have described prevalence rates of CVD risk factors in PLHIV on ART, our study described the metabolic profiles of ART-naïve Ugandan PLHIV at ART initiation, thus offering an opportunity to improve prevention strategies in a population with a high CVD risk.

## CONCLUSION

We demonstrated that despite being a relatively young population, cardiometabolic risk factors were common among Ugandan PLHIV initiating DTG-based ART, underscoring the need for programmatic implementation of surveillance and management of NCD comorbidities among PLHIV initiating ART in Uganda and similar settings.

## Data Availability

All data produced in the present work are contained in the manuscript

## DECLARATIONS

### Ethics approval and consent to participate

Ethical approval to perform the study was obtained from The AIDS Support Organization (TASO) Institutional Review Board (registration number: TASOREC/051/2020-UG-REC-009) and the Uganda National Council for Science and technology (Registration number: HS1032ES). All participants signed informed consent prior to enrolment in the study.

### Availability of data and materials

The datasets used and/or analyzed during the current study are available from the corresponding author upon reasonable request.

### Potential Conflicts of Interest

All authors declare no competing interests

### Funding

The study received funding support from NIH-Fogarty University of California (UCSF) Global Health Fellowship Program, grant number: 2D43TW009343-06 (F.M., J-M.S.), the National Institutes of Health, UCSF-Gladstone Center for AIDS Research, grant number: P30AI027763 (F.M., J-M.S.) and the Fogarty International Center of the National Institutes of Health, Award Number D43TW009771 (B.C.). Additional funding were provided through the Roe Green Center for Travel Medicine and Global Health/University Hospitals Cleveland Medical Center (W.A., G.A.Y.).

### Authors’ contributions

Conceptualization: W.A., F.M., G.A.Y. Methodology: W.A., F.M., G.A.Y. Resources: F.M., B.C., N.B., S.D., M.E., J-M.S. Formal analysis: G.A.Y. Investigation: W.A., F.M., M.E., S.D. Visualization: W.A., F.M., G.A.Y. Writing–original draft preparation: W.A., F.M., G.A.Y. Writing–review and editing: W.A., F.M., B.C., N.B., J-M.S., M.E., S.D., R.A.S., G.A.Y. Funding acquisition: W.A., F.M., B.C., N.B., J-M.S., G.A.Y. All authors contributed important intellectual content and participated in the interpretation of results, writing and editing of the final manuscript draft.

## Acknowledgments

We acknowledge the Makerere University Infectious Diseases Institute and the Kampala City Council Authority (KCCA) for permitting us to use the Kisenyi HIV clinic for the study. we also acknowledge the administrative support from the Kisenyi Health Center IV administration. Lastly, we acknowledge the patients that participated in our study.

## Notes

### Competing Interest Statement

The authors have declared no competing interest.

